# Evidence Based Gait Analysis Interpretation Tools (EB-GAIT)

**DOI:** 10.1101/2025.02.19.25321739

**Authors:** Michael H. Schwartz, Andrew G. Georgiadis

**Affiliations:** Orthopedic Surgery, University of Minnesota - Twin Cities, Minneapolis, USA; Center for Gait and Motion Analysis, Gillette Children’s Specialty Healthcare, St. Paul, USA

## Abstract

Clinical gait analysis (CGA) has historically relied on clinician experience and judgment, leading to modest, stagnant, and unpredictable outcomes. This paper introduces *Evidence-Based Gait Analysis Interpretation Tools* (EB-GAIT), a novel framework leveraging machine learning to support treatment decisions. The core of EB-GAIT consists of two key components: (1) *treatment recommendation models*, which are propensity models that estimate the probability of specific surgeries based on historical standard-of-practice (SOP), and (2) *treatment outcome models*, which predict changes in patient characteristics following treatment or natural history.

Using Bayesian Additive Regression Trees (BART), we developed and validated treatment recommendation models for 12 common surgeries that account for more than 95% of the surgery recorded in our CGA center’s database. These models demonstrated high balanced accuracy, sensitivity, and specificity. We used Shapley values for the models to enhances interpretability and allow clinicians and patients to understand the factors driving treatment recommendations. We also developed treatment outcome models for over 20 common outcome measures. These models were found to be unbiased, with reliable prediction intervals and accuracy comparable to experimental measurement error.

We illustrated the application of EB-GAIT through a case study, showcasing its utility in providing treatment recommendations and outcome predictions. We then use simulations to show that combining propensity and outcome models offers the possibility to improve outcomes for treated limbs, maintain outcomes for untreated limbs, and reduce the number of surgeries performed. The EB-GAIT approach addresses the limitations of the conventional CGA interpretation method, offering a more structured and data-driven decision-making process.

EB-GAIT is not intended to replace clinical judgment but to supplement it, providing clinicians with a second opinion grounded in historical data and predictive analytics. While the models perform well, their effectiveness is constrained by historical variability in treatment decisions and the inherent complexity of patient outcomes. Future efforts should focus on refining model inputs, incorporating surgical details, and pooling data from multiple centers to improve generalizability.

EB-GAIT represents a significant step toward precision medicine in CGA, offering a promising tool to enhance treatment outcomes and patient care.

**Plain Language Summary:** Clinical gait analysis (CGA) helps doctors understand walking problems in patients with movement disorders, like cerebral palsy. Traditionally, treatment decisions have relied on doctors’ experience and judgment, which can lead to inconsistent and unpredictable outcomes. To address this, we developed *Evidence-Based Gait Analysis Interpretation Tools* (EB-GAIT), a new method that uses advanced computer models to guide treatment decisions.

*What is EB-GAIT?:* EB-GAIT combines two types of models: (1) *Treatment Recommendation Models* that predict the likelihood of specific surgeries based on historical treatment patterns at our center, and (2) *Outcome Prediction Models* that estimate how patients’ orthopedic deformity, walking pattern, and mobility might improve after surgery or without treatment. Using a powerful statistical tool called Bayesian Additive Regression Trees (BART), we built models for 12 common surgeries and over 20 outcome measures. These models are accurate, unbiased, and provide reliable predictions.

*How Does EB-GAIT Help?:* EB-GAIT helps doctors understand which treatments are likely to work best for each patient. The models explain why a treatment is recommended, making it easier for doctors and patients to understand. Simulations show that combining treatment recommendations and outcome predictions can lead to better results for patients, fewer unnecessary surgeries, and more consistent care.

*A Case Study:* We use a case study throughout the paper to demonstrate EB-GAIT using a real patient example. The models accurately predicted the outcomes of surgery, showing how EB-GAIT can support doctors in making evidence-based decisions.

*The Future of EB-GAIT:* While EB-GAIT is a powerful tool, it doesn’t replace doctors’ expertise. Instead, it provides additional information to help doctors and patients make informed choices. In the future, we plan to improve the models by adding more details about surgeries and combining data from multiple centers. We view EB-GAIT as a big step forward in using data and technology to improve care for patients with walking problems. By making treatment decisions more precise and personalized, EB-GAIT aims to help patients move better and live fuller lives.

## 1. Background

Historically, treatment decisions in CGA have relied on clinicians’ collective experiences and judgments, which may not be consistently evidence-based nor optimal. Outcomes resulting from this *ad hoc* approach have been modest, stagnant, and unpredictable. This manuscript presents a novel approach for using machine learning models to inform treatment decisions in clinical gait analysis (CGA). We call these “*evidence-based gait analysis interpretation tools*” — or EB-GAIT. The EB-GAIT framework consists of two key components: (1) treatment recommendation models, which estimate the probability of a limb receiving specific surgeries based on historical standard-of-practice (SOP), and (2) outcome models, which predict changes in patient characteristics either following surgery or reflecting their untreated natural history.

This manuscript is intended for clinicians and bioengineers involved in CGA, where extensive data sources — including three-dimensional instrumented motion analysis — guide treatment for individuals with movement impairments. The innovation lies not in the machine learning techniques — which are well-established — but in their application to clinical practice.

### 1.1 The Conventional Approach to Interpreting Clinical Gait Data

The process of incorporating CGA data into treatment decisions for patients is an earnest but unstructured meeting of the minds among experts. We will refer to this process as the “*conventional approach*” to data interpretation. In the conventional approach, large amounts of complex data are presented for a patient with an incompletely understood and often incurable condition. These data are then discussed among clinical and biomechanical experts using experience, recall, and logic as a basis for understanding the numbers, graphs, and images in front of them. At many centers, including ours, the interpretation is often led by a physician who is unfamiliar with the patient, and has never seen or examined them in person. These experts then decide on a set of treatment recommendations. The conventional approach has been shown to exhibit only moderate interobserver reliability between centers, though more consistent results have been demonstrated within a single center [2].

### 1.2 The Historical Standard of Practice and Outcomes

The output of the conventional approach is a set of treatments, which, over time, form the historical standard of practice (SOP) at a particular center. A consistent SOP does not imply that the interpretation is evidence-based or leads to optimal outcomes. We have previously shown that elements of the SOP — such as recommending femoral derotation osteotomies for patients presenting with excessive femoral anteversion by examination but typical transverse plane foot progression — lead to clearly sub-optimal outcomes [3]. Despite the earnest effort of the clinicians, historical outcomes across a range of global measures are modest in magnitude, stagnant over time, and unpredictable for individual patients [Figure 1].

**Figure 1.**
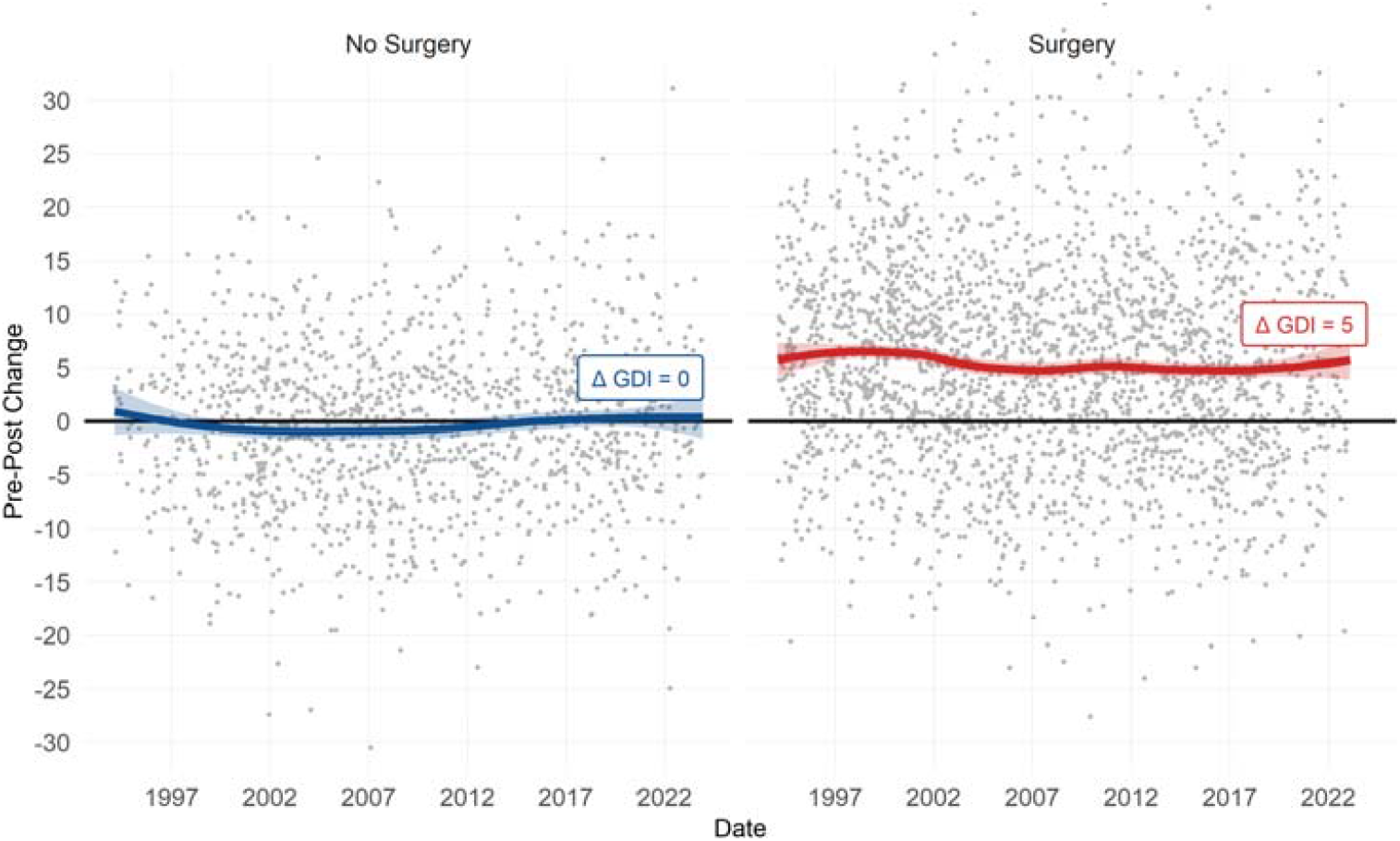
Historical outcomes after major orthopedic surgery are modest, stagnant, and unpredictable. Here we show this for overall kinematic outcomes (Gait Deviation Index, GDI)-though the results are similar for every outcome measure we have examined. Each point is the limb of an individual. The solid line is a LOESS smoothing curve (span = 0.7).

### 1.3 Machine Learning to Establish an Evidence-Based Standard of Practice

Machine learning models trained with clinical gait data can predict treatments and outcomes [3–10]. In this manuscript we describe a method for using the output from such models to guide treatment decisions. The manuscript is organized into three main topics: (1) Treatment Recommendation Models: We build and test models that capture the historic SOP for assigning a treatment, (2) Treatment Outcome Models: We build and test models for estimating changes in patient characteristics with treatment or due to untreated natural history, and (3) Benefit of Using Treatment Recommendation and Outcome Models: we estimate the future benefit of implementing the EB-GAIT approach in practice.

## 2. Methods

### 2.1 Ethical Compliance

This study was reviewed and authorized by the University of Minnesota institutional review board review (STUDY00012420). All experiments were performed in accordance with relevant guidelines and regulations. Written consent for use of medical records was obtained at the time of service from all participants or their legal guardian. An option to rescind this permission was offered to patients at every visit to our center. Data was accessed on May 25, 2024. Authors did not access information that could identify individual participants during or after data collection.

### 2.2 Treatment Recommendation Models

In this section, we describe the methodology for developing, testing, interpreting, and implementing treatment recommendation models. These models provide evidence-based guidance by estimating the likelihood of specific surgeries based on the historical SOP at our center. The term *propensity model* is a general and widely accepted descriptor for models of this kind. Throughout this manuscript, we use the terms *propensity model* and *treatment recommendation model* interchangeably.

#### What is a Propensity Model?

“*Propensity model*” is a broad term describing a set of computations that estimate the probability of an event. We will use propensity models as the tool for making treatment recommendations. Here, the event in question is a *limb* undergoing a specific surgery based on the historic SOP at *our center*. We write a propensity model as:

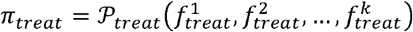

Here π_*treat*_ is the probability a limb will be prescribed the treatment treat, the function P_*treat*_ is a set of “*rules*”, and 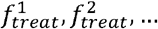 are clinically relevant variables (“*features*”) specific to the treatment being considered, like ankle range-of-motion, age, dynamic equinus, and so on.

We focus on twelve common surgical procedures that collectively account for over 95% of the surgeries documented in the clinical database of our gait center. These surgeries are: selective dorsal rhizotomy, rectus femoris transfer, psoas lengthening, hamstrings lengthening, adductor lengthening, calf muscle lengthening, femoral derotation osteotomy, tibial derotation osteotomy, patellar tendon advancement, distal femoral extension osteotomy, foot and ankle bony reconstruction (*e*.*g*., calcaneal osteotomy, talo-navicular fusion), foot and ankle soft tissue reconstruction (*e*.*g*., posterior tibialis tendon transfer, split tibialis anterior tendon transfer). The approach we present is not limited to these surgeries. Any treatment — surgical or otherwise — for which there is sufficient baseline and follow up data can be evaluated.

#### Building and Testing Propensity Models

A panel of clinical experts selected a set of features for each of the twelve surgeries analyzed in this study (Appendix 1). The selection process relied on the panelists’ collective expertise and clinical judgment, prioritizing both model parsimony and clinical interpretability. It is important to acknowledge that the selected features may not be optimal, and future refinements — such as adding, removing, or reweighting features — could potentially improve model performance. Feature selection is a critical component of machine learning model development, and extensive research exists on this topic, which lies beyond the scope of this manuscript.

Once the features were chosen, we fit Bayesian Additive Regression Trees (BART) models for each surgery [11]. Other modeling frameworks can be used, ranging from logistic regression to random forests. The purpose of this manuscript is not to explain or promote BART, which is a well-understood standard tool. However, it may be useful for the reader to understand practical reasons why we chose BART as modeling framework. Disinterested readers can skip the following discussion of BART without affecting their ability to understand the rest of this manuscript.

#### Why BART?

##### BART is able to model complex response surfaces

This includes nonlinearities, bifurcations, and so on. These are common in real-world treatment decisions. An example of a nonlinearity is the effect of tibial torsion on the decision to perform a tibial derotational osteotomy [Figure 2]. Within the typical torsion range, there is almost no impact of torsion on the decision to derotate. As torsion deviates external or internal to the typical range, the probability of a derotation rises. There are many other situations the reader can imagine where nonlinearities exist. Largely because of its flexibility in modeling complex responses, BART has been shown to be a class-leading causal modeling framework [12].

**Figure 2.**
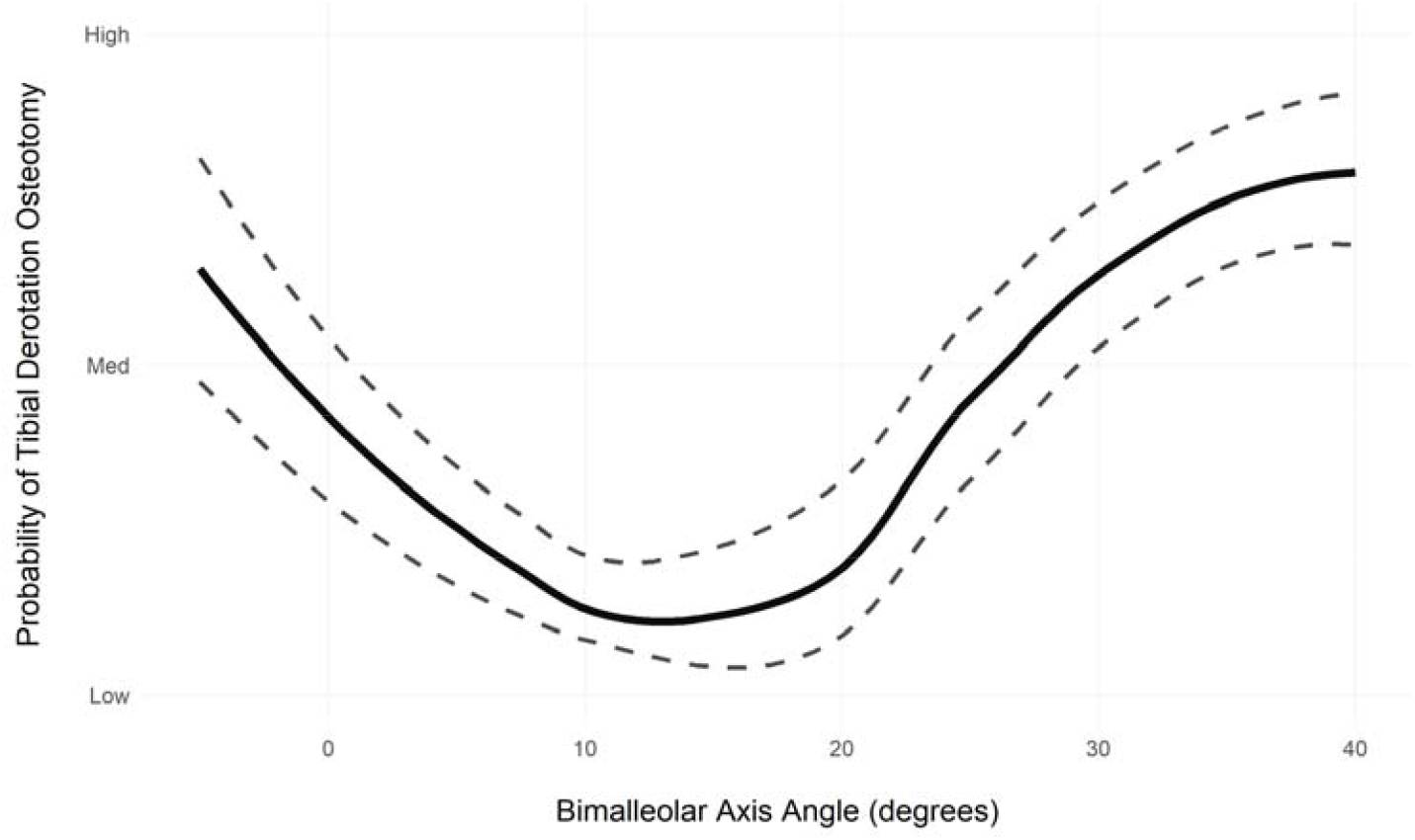
Effect of tibial torsion (bimalleolar axis angle) on the probability of a limb undergoing a tibial derotation osteotomy. The response is highly nonlinear. Close to typical values (∼15 degrees) tibial torsion has little effect. As internal (smaller) or external (larger) torsional deviations appear, the probability of surgery rises. A linear propensity model, like logistic regression, would not be able to capture this response.

##### BART is able to handle missing data with minimal assumptions

There is no imputation needed and BART treats missingness itself as a feature [13]. This reduces concerns about biases introduced from data that is missing in a systematic manner, such as missing ROM measurements on severely contracted limbs, or missing strength measurements on patients who cannot understand or follow directions. By handling missing data in this principled and user-friendly fashion, BART allows applications of the model to patients with missing data. This is a common, and often non-random occurrence. For example, patients with more severe impairments may not be able to complete or cooperate with instructions, leading to missing data. Excluding these patients would introduce important biases. The ability to competently handle missing data also promotes use of models in centers that may not collect all the measurements described in this study.

##### BART is Bayesian

As a Bayesian model, BART outputs a probability distribution of results — called a posterior. This allows the uncertainty of any prediction to be observed and reported directly. The Bayesian formulation also provides for built-in regularization — meaning that BART models do not “*overreact*” to individual features. An important consequence is that BART models generally produce nearly optimal results without the need for manual hyperparameter tuning. This is a stark contrast to many advanced modeling approaches.

##### BART is fast, free, and easy to use

There are several packages that implement BART in the `R` programming language. We use the `bartMachine` package [14]. There is vigorous ongoing innovation and development related to BART.

#### Fitting Propensity Models

We evaluated 7546 gait assessments from 2090 individuals referred to our center by 176 different clinicians. The individuals all had baseline and follow-up CGA visits within a three-year span. We lumped diagnoses into 10 categories. Due to our referral patterns, individuals diagnosed with cerebral palsy dominated the data set (84% cerebral palsy). The mean (sd) baseline age was 8.9 (3.3) years and there were 42% females by sex at birth.

When the treatment recommendation model is a BART model, then treatment probability is a posterior distribution of probabilities (posterior) and the probability of prescribing the treatment is the mean of this posterior. The treatment categories (recommended or not) are defined by the mean probability compared to a critical threshold, which is 0.5 by default.

As is typical in a clinical service, we have an imbalanced data set, with many more untreated limbs than treated. For example, out of 7546 limbs in the database, only 334 of them underwent an adductor release. This makes evaluating propensity model performance challenging. Consider that for an adductor release, with a prevalence of only 4%, a model that predicts no limb gets an adductor release would be 96% accurate. To overcome this challenge, we begin by recognizing that we want a model with good balanced accuracy, which implies good sensitivity and specificity. Alternatives choices would include fitting a model with good accuracy, or high negative predictive value, or high Brier score, etc. By focusing on balanced accuracy, we are implicitly demanding that “*If the historical SOP resulted in treatment, we want the propensity model to recommend treatment. If the historical SOP resulted in no treatment, we want the propensity model to recommend no treatment*.”

We use a standard approach for building a propensity model with imbalanced data. First, we split the dataset into training and testing sets (70% and 30%, respectively). Then we choose the treatment of interest. Next we use under-sampling to generate a balanced sub-sample of the training set with equal numbers of treated and untreated (control) limbs [15]. Finally, we use the balanced sub-sample to fit a BART model to compute the probability that the limb underwent the selected treatment. Finally, we test the model performance on the independent test set. Note that the test set is *not* balanced through undersampling, and therefore exhibits the same imbalanced surgical prevalences that are observed in practice. Using an unbalanced test set gives a realistic estimate of model performance on future clinical data.

#### Propensity Model Performance

We report standard classifier metrics to give a clear picture of how well the classifier will work in practice [Table 1]. Examining the performance, we can make several general statements about the propensity models. Balanced accuracy is consistently high, ranging from the mid .70s to .90. Specificity and sensitivity are consistently high and balanced, ranging from 0.71 to 0.92. The mean specificity and sensitivity were 0.77 and 0.78, respectively. Note that sensitivity and specificity were even high for low prevalence surgeries with known low inter-surgeon consistency, such as psoas lengthening. Also, keep in mind that the upper limit of propensity model performance is governed by how consistently clinicians made treatment decisions in the past. Treatment decisions on the same data are known to vary widely between clinicians and institutions [1]. Model performance was strong even for the generic “*Foot and Ankle …*” categories, which are broad and based on almost entirely subjective assessment of foot deformity.

**Table 1:**
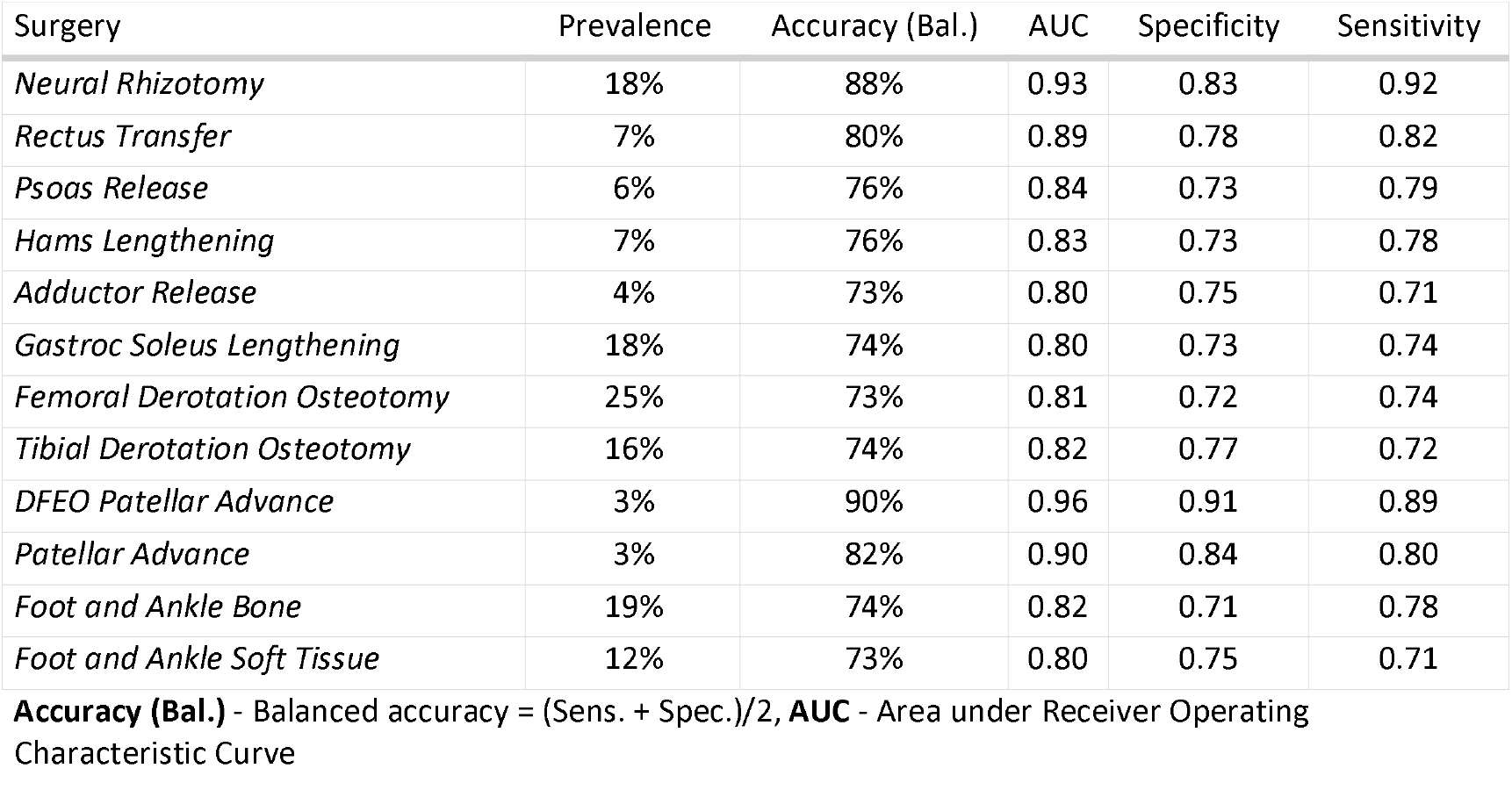
Propensity Model Performance.

We can put the propensity models’ performance in context by examining the specificity and sensitivity estimates for several important and common medical screening tests: Mammography to detect breast cancer exhibits specificity .90 and sensitivity .54 -.94 [16]. Colonoscopy to detect adenomas 6 mm or larger exhibits specificity ∼.90 and sensitivity from .73 - .98 [17]. Pap smears to detect cervical cancer exhibit specificity ∼.75 and sensitivity ∼.68 [18]. Rapid antigen tests for COVID-19 exhibit specificity ∼.99 and sensitivity ∼.65 (.44 in asymptomatic individuals) [19]. Finally, prostate specific antigen testing to detect prostate cancer exhibits specificity ∼.90, sensitivity from .09-.33 [20].

#### Propensity Profile

We will be referring to a case study throughout this manuscript to give an idea of how EB-GAIT looks and is used in practice. All protected health information has been removed or replaced. Some variables — like age — have been altered slightly to preserve the general sense of the individual while maintaining anonymity. The case was selected from a search of our database for individuals who underwent calf muscle lengthening consistent with the propensity model and had a follow-up visit at around one year post-surgery. The chosen patient had birth, developmental, and treatment history typical for someone diagnosed with hemiplegic cerebral palsy. On the right side, during the same surgical event that included the calf muscle lengthening, the individual also underwent a femoral derotation osteotomy, hamstrings lengthening, and bony foot reconstruction. It may be interesting for readers to know that, when choosing this case, we looked at the follow-up video — so we had a sense of the outcome — but we *did not* look at the propensity or outcome model results to see how accurate they were. Only one other patient was screened before selecting this patient. We mention all of this to give the reader some indication of the degree of “*cherry-picking*” involved in the case-study selection. We are deeply skeptical of case-studies due to the biases they often reflect [21]. A deidentified version of the full EB-GAIT report for this case study is provided as supplementary material for this manuscript.

In practice, we provide a compact display of the surgery recommendations for 12 surgeries. Below is the propensity profile for the case study [Figure 3]. The categories are divided into probability quintiles and color-coded from high probability (green) to low probability (purple). This color-blind-safe scheme for propensity will be maintained in several other places to show indications (green) and counterindications (purple) for surgery. It will also be used to indicate outcome predictions for treated (green) and untreated (purple) limbs.

**Figure 3.**
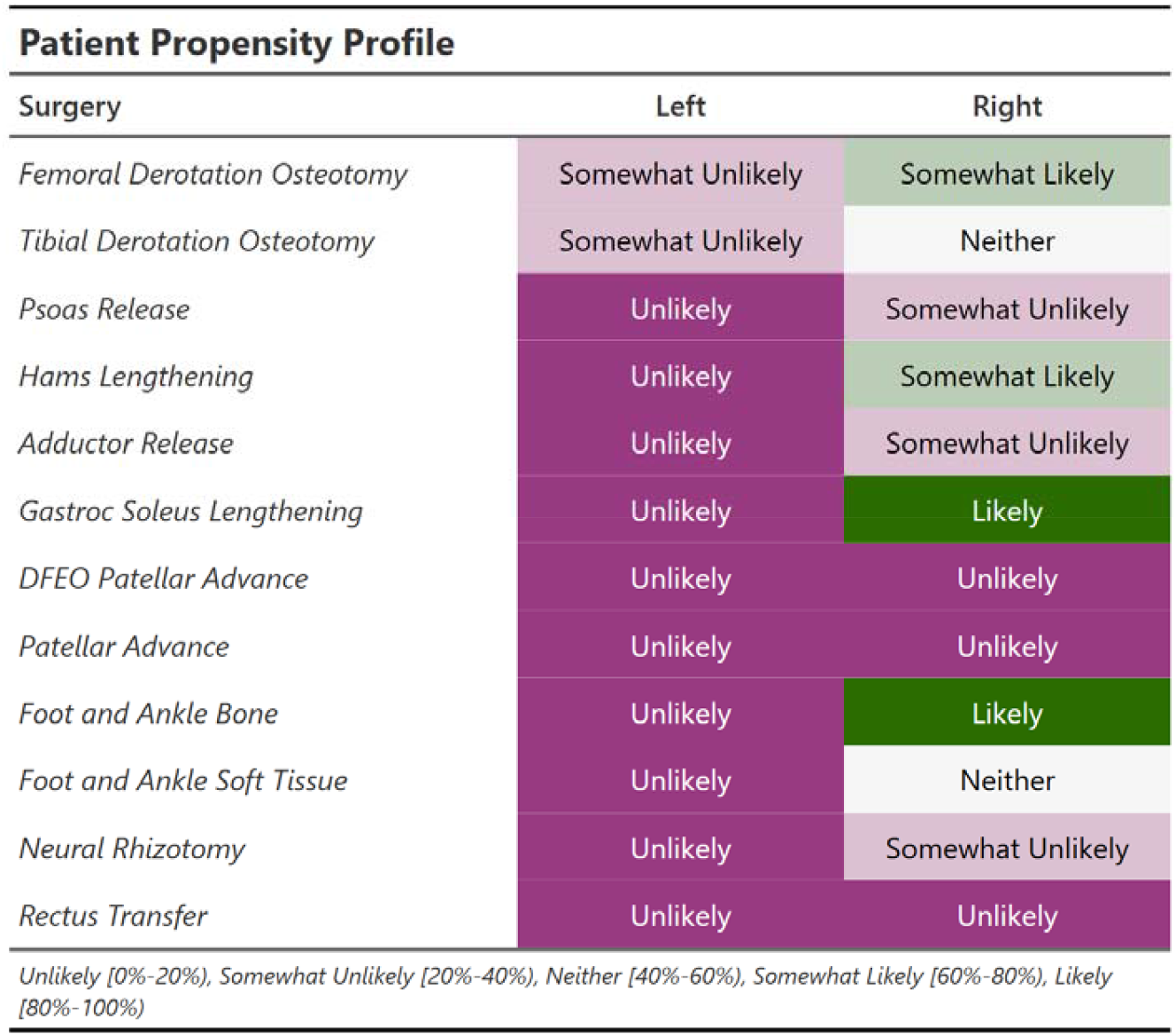
Propensities for 12 surgeries. Probability of surgery is categorized and color coded for quick reference.

#### Propensity and Outcomes

Given the skill and experience of the surgeons whose practices shape the SOP, it is reasonable to expect that adherence to the SOP would yield consistently positive outcomes. However, as previously demonstrated [Figure 1], historical outcomes have been modest, stagnant, and unpredictable. It is important to recognize that historical outcomes reflect actual clinical decisions, which do not always align with the SOP. There are many legitimate reasons why the SOP may not have been followed, including the fact that the SOP is not formally documented, as well as the absence of critical patient- or clinician-specific factors such as patient goals, family experiences with prior treatments, perceived ability of the patient to engage in rehabilitation, and so on.

The variability in adherence to the SOP provides a unique natural experiment, allowing us to investigate whether following the SOP is leads to improved outcomes. To explore this, we calculated the concordance between administered treatments and the SOP (as estimated by the propensity models) and examined how outcomes varied with concordance. High concordance indicates that treatments closely followed the SOP, whereas low concordance reflected deviations from the SOP.

We compute concordance for each of the 12 surgeries (*K*_*treat*_) as follows. First, we rescale surgery probability from -1 to 1:

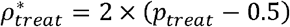

Next, we square the scaled surgery probability to emphasize high and low probabilities while preserving the sign,

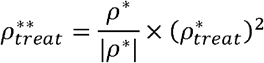

This transformation amplifies differences at the extremes while compressing moderate probabilities. Concordance is then defined as 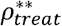 if surgery was administered, and 0 otherwise. This approach rewards surgeries performed on limbs with high predicted probabilities, penalizes those performed on limbs with low probabilities, and remains neutral when no surgery is performed or when the predicted probability is near 50%. The *overall* concordance is the sum of the 12 individual concordances:

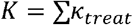

We demonstrate the effect of concordance with SOP on outcomes by examining calf muscle lengthening. We first compute the concordance for calf muscle lengthening as described above (*κ*_*CalfLen*_). Next we choose relevant outcomes at the body structures level (*e*.*g*., Maximum Ankle Dorsiflexion (Knee Extended)) and at the body functions level (*e*.*g*., Initial Contact Ankle Angle Sagittal Plane). Finally, we examine the change in these outcome measures after calf lengthening surgery as a function*κ*_*CalfLen*_ [Figure 4]. There is a clear and strong benefit to following the historical SOP — though the variation around the trend is wide. Other surgeries and outcomes exhibit similar responses.

**Figure 4.**
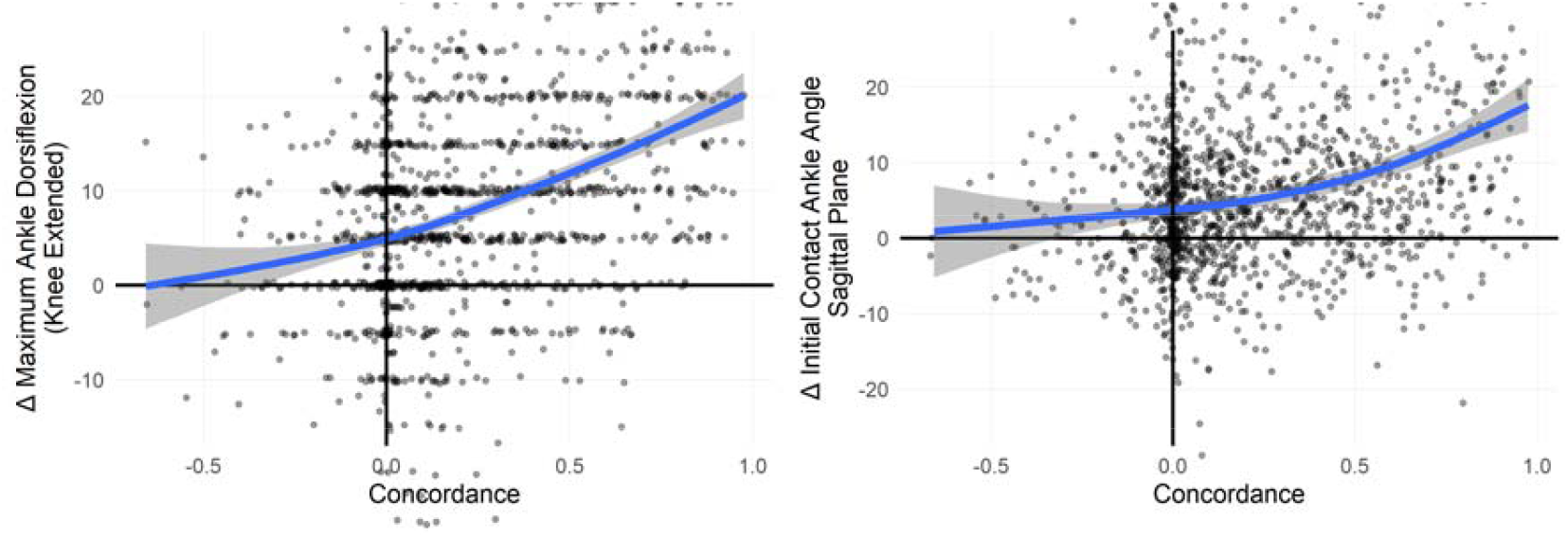
Calf lengthening surgery on limbs that reflect the historical SOP (high concordance) results in large improvements in both static ankle range of motion and dynamic ankle dorsiflexion in gait. Surgery that does not conform to the SOP leaves these measures unchanged, or even worsened.

A similar analysis can be conducted at an global level by evaluating the gait kinematic response (*Δ*GDI) to overall concordance *K* [Figure 5]. As with the isolated surgery results, there is a clear message that following the historical SOP is generally beneficial, although there is significant variability around the mean response.

**Figure 5.**
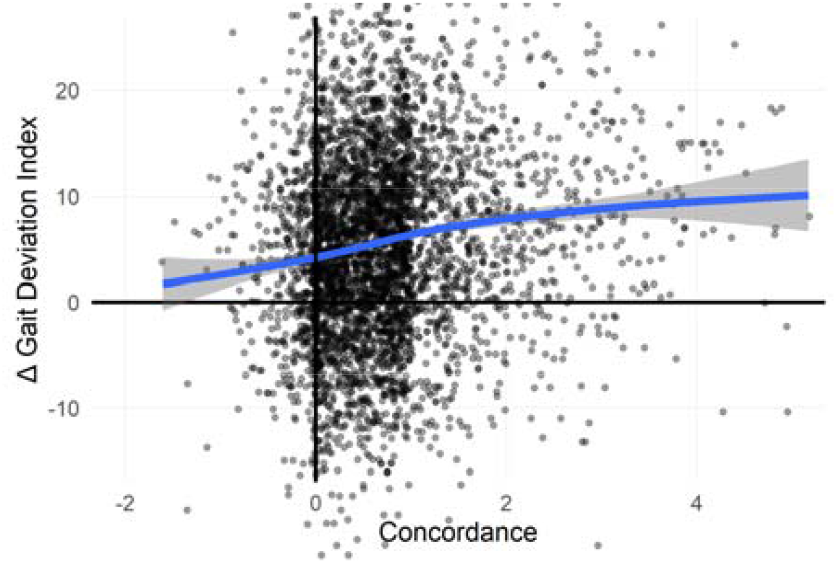
Following rather than flouting the historical SOP leads to better GDI outcomes for treated limbs. For example, the change in GDI at an overall concordance of 4 is approximately 10 points, whereas at a concordance of -2 it is approximately 0.

#### Understanding the Propensity Models

##### Understanding at an Overall Level - Partial Dependence Plots

The BART models that compute the propensity scores are “*black boxes*”. The input features for the models are clinically relevant, but it is not easy to discern how the BART model computes the surgery probability based on these features. The actual calculation involves summing the results of many shallow decision trees constructed using Bayesian principles. There are a number of standard tools that can be used for interrogating models like this. The use of a post-processing technique to give a glimpse at the inner workings of a complex machine learning model is called “*explainable machine learning*”. Note that there are machine learning experts who advocate for simpler models that do not require an explanation, and are instead interpretable in their raw form [22].

At the simplest level we can examine the marginal effect of an individual feature, averaged across values of all other features. This is called a partial dependence plot [23]. Partial dependence plots produce an overview of features’ impact. However, partial dependence plots ignore interactions. For example, restricted ankle range of motion may be an indication for calf muscle lengthening, but the effect may vary with patient age. The interaction of age and ankle ROM on calf lengthening probability is not captured by a partial dependence plot. Furthermore, partial dependence plots average effects across conditions that may be unrealistic. For example, the partial dependence plot for age averages the effect across all heights — so the effect of age = 6 may include a hypothetical 6-year-old who is 2m tall. As such, partial dependence plots can be considered *contextless*. Nevertheless, partial dependence plots are a standard tool for explaining models such as BART.

##### Example: Rectus Femoris Transfer

Consider the partial dependence plot for rectus femoris transfer [Figure 6]. We describe some, but not all, of the sub-plots. *Age*: The probability of surgery rises steadily until around age 10, then plateaus. *Maximum Swing Knee Angle Sagittal Plane (Max. Knee Flexion)*: The probability of surgery is high until just over 50^°^ peak knee flexion, steadily decreases until around 60^°^, then plateaus. *ROM Swing Knee Angle Sagittal Plane (Knee Flexion ROM)*: The probability of surgery is high for limited ROM, decreasing slowly until around 35^°^, after which it decreases rapidly. *Era*: The probability of rectus transfer surgery has steadily decreased over time. *Rectus Femoris Spasticity*: The probability of surgery increases with increasing spasticity up to modified Ashworth scores of 2, then declines slightly for scores of 3 and 4. *Prior Rectus Femoris Transfer*: A prior rectus femoris transfer makes a repeat surgery less likely.

**Figure 6.**
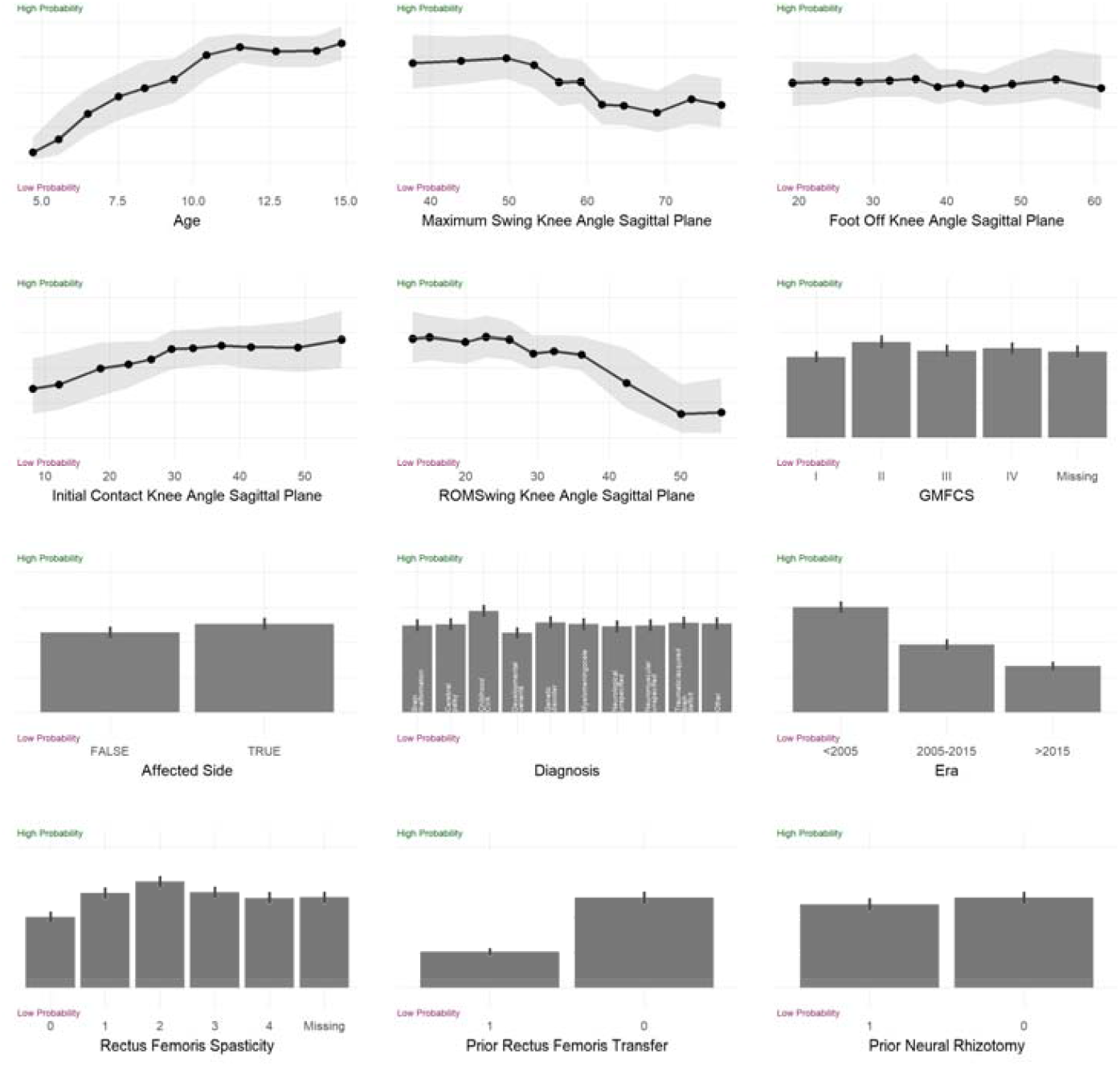
Partial dependence plot for the rectus femoris transfer propensity model.

##### Understanding at an Individual Level - Shapley Values

In practice, we seek to understand the influence of a specific feature on an individual prediction — for example, how much a patient’s knee flexion ROM contributes to their probability of being prescribed a rectus femoris transfer. Partial dependence plots do not provide this level of insight. Instead, Shapley values offer a rigorous, principled approach to quantifying feature contributions [24]. The Shapley value for a feature represents its contribution to a limb’s surgery probability, relative to the overall mean probability observed in the model’s training data. Since Shapley values are computed with respect to the mean probability of a surgery, they depend on the local SOP and will vary across clinical centers.

For example, suppose the overall probability of a rectus transfer is 15% overall. Consider *patient X*, whose left knee flexion range-of-motion has a Shapley value of -11%, while their age has a Shapley value of +6%. Assuming all other Shapley values are zero, the probability of *patient X* undergoing a rectus transfer, based on the historical SOP, would be:

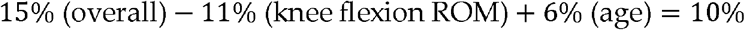

Formally, the Shapley values for the propensity models estimating the SOP-based probability of undergoing treat satisfy the following additive relationship:

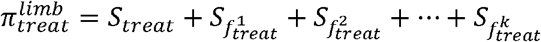

Where 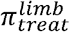 is the probability that a limb undergoes the treatment, *s*_*treat*_ is the mean probability of treat across all limbs in the training set, and 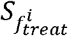 is the Shapley value for the *i*^th^ feature for the propensity model predicting treat.

Thus, the 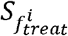 are contributions from features such as age, diagnosis, and so on(s_*age*_, *s*_*diagnosis*_, …). Since s_*treat*_ is the same for all limbs, only the 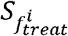 values determine individual differences in treatment probability.

A positive 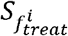 indicates that feature 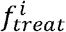 increases the likelihood of undergoing treat. We call this an indication. Conversely, a negative 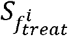 signifies a *counter-indication*, meaning the feature reduces the likelihood of treatment. The magnitude of 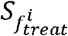 reflects the strength of the indication or counter-indication.

We can compute Shapley values for a limb and present the results in a compact tabular format, sorted from strongest indications to strongest counter-indications for ease of interpretation. Below we show the Shapley values for calf muscle lengthening (Gastroc-Soleus Lengthing) for the case study [Figure 7]. We only show the feature-specific Shapley values, 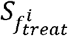, excluding the mean Shapley value.

**Figure 7.**
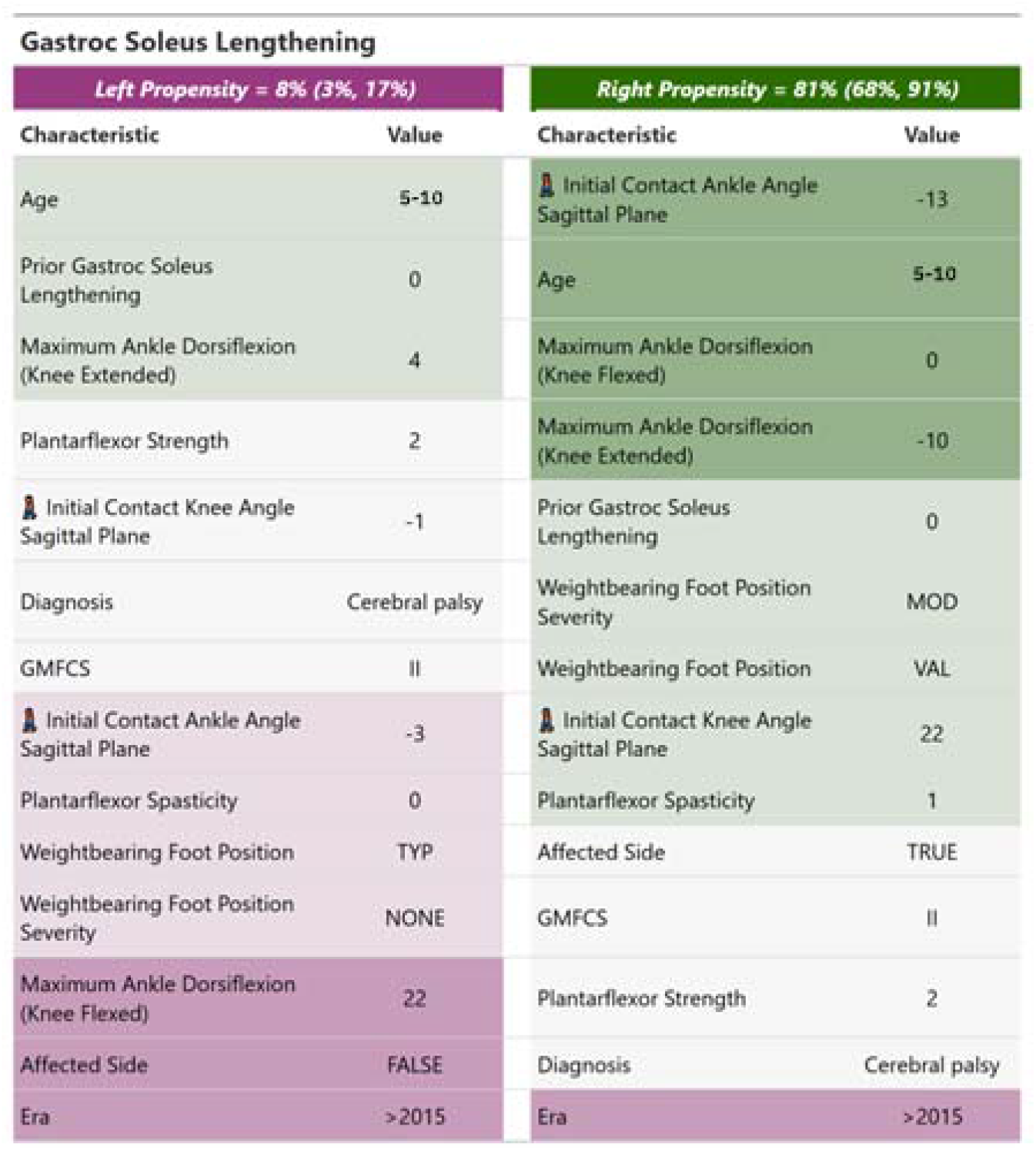
“Shapley values presented in a tabular form. Columns correspond to sides. The features are sorted and color-coded from strongest indication to strongest counterindication. The value of the feature is shown. An emoji of a walking person indicates a feature extracted from instrumented gait analysis (as opposed to physical examination). The probability for surgery is indicated at the top of each column.”

For the case study, the right limb exhibits a high probability for calf muscle lengthening surgery. The color-coding of indications and counterindications scales with magnitude. The indications (green) for treatment are initial contact ankle angle in the sagittal plane (from the instrumented gait data — indicated by an icon of a walking person), age, maximum ankle dorsiflexion (from the physical examination). There are also several weak indications (light green) and only one counterindication (purple), which is our center’s trend towards less calf muscle lengthening surgery over time. On the left (unaffected) side, surgery is unlikely, and there are numerous counterindications, including the fact that this is the unaffected side of a patient with a diagnosis of hemiplegia.

The probability of surgery for the limb, shown in the top row of Figure 7, is represented as a range rather than a single value. This range corresponds to the 90% prediction interval derived from the Bayesian posterior of the propensity model. The uncertainty in the surgery probability arises from several factors, such as model imperfections, historical variability in decision-making, measurement errors, and the limited size of the dataset used to train the model. The unaffected side displays a relatively narrow range of propensity uncertainty, whereas the affected side shows a broader range — though it remains entirely above the 50% likelihood threshold.

### 2.3 Outcome Prediction Models

We have shown that propensity models for treatment recommendations can reliably identify the historical SOP. We also showed that adherence to the SOP is generally associated with favorable outcomes [Figure 4, Figure 5]. However, substantial variability in outcomes persists, even when the SOP is followed.

Additionally, propensity models — and the data they rely on — have inherent limitations, preventing them from capturing all nuanced and critical factors influencing surgical decisions. These considerations highlight the need for a broader decision-making framework. Simply put: “*Propensity is not fate!*”

From a Bayesian perspective, propensity models provide valuable evidence that clinicians can use to update their prior beliefs about the treatments that may be appropriate for the patient under evaluation. These beliefs can be further refined by incorporating additional information in the form of treatment outcome estimates. In the following section, we develop and evaluate models designed to provide this critical supporting information.

#### What is an Outcome Prediction Model?

An treatment outcome model is a set of rules of the form:

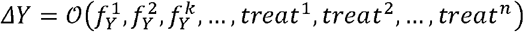

Where *Y* is a chosen variable, 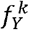 are clinical features deemed relevant to changes in *Y*, and(*treat*^1-*n*^) are proposed treatments. Note that *treat*^*i*^ = 0 if the treatment does not occur and *treat*^*i*^ = 1 if it does.

#### Fitting Outcome Prediction Models

We again used BART as our framework for building treatment outcome models, employing a 70%-30% training-testing data split. A panel of clinical experts selected a number of important outcome variables (*Y*) at the levels of body structures and body functions. These variables were chosen based on clinical relevance to the 12 surgeries (*treat*^*i*^) for which we fit treatment recommendation models. For example, changes in femoral torsion and mean stance hip rotation during gait were deemed relevant to a femoral derotation osteotomy, while changes in ankle dorsiflexion ROM and initial contact dorsiflexion during gait were relevant to calf muscle lengthening. We also built models for predicting changes in gait kinematics (Gait Deviation Index) and mobility (Functional Assessment Questionnaire Transform). Currently, we have models for around 20 different outcome variables, with additional outcome models built and tested as needed.

For each model, the same clinical experts chose the predictive features that were felt to be relevant to the treatment outcome. The features consisted of the baseline value of the outcome variable, relevant structural and biomechanical variables, and whether the limb underwent each of the 12 major surgeries of interest. The interval and prior treatment status for each surgery was dichotomous, and did not include any information about dose (*e*.*g*., amount of derotation) or technique (*e*.*g*., Baker vs. Strayer calf muscle lengthening). The complete list of features for each outcome are listed in Appendix 2.

We used an *ad hoc* iteration of features to fit models that were accurate and parsimonious. However, there is no guarantee that the current models are optimal. A substantial body of research exists on optimal feature selection for machine learning, and future efforts will include improving the models. Despite the lack of optimization, the models are well-calibrated and provide useful precision, as discussed below.

#### Outcome Prediction Model Performance

Clinically useful models must have reasonable accuracy, minimal bias, and reliable precision. An unbiased model ensures that its predictions, on average, align closely with actual observed values, without systematic over- or underestimation. Reliable precision refers to the validity of the model’s prediction intervals (PIs), which should consistently encapsulate the stated proportion of future outcomes. For instance, a 90% PI should contain 90% of the observed values in prospective applications. The outcome models, like the propensity models, were evaluated retrospectively, using independent test data.

The treatment outcome models performed exceptionally well in terms of low bias and reliable precision. Here we report performance for a representative subset of the treatment outcome models at body structures and body functions levels [Table 2]. Accuracy, as measured by mean absolute error (MAE) varied with outcome. For example, the MAE for *Δ* ankle dorsiflexion ROM was around 6^°^, which is similar to the test-retest measurement error (∼ 8^°^). A similarly strong performance was seen across all physical examination measures. At the kinematic level, MAEs generally reflected a slightly larger fraction of test-retest measurement error. One explanation for these larger errors is that the effects of treatment on causally downstream outcomes is more complex, and thus more difficult to predict. For example, the effect of a femoral derotation osteotomy on femoral torsion (body structure) is reasonably straightforward given the nature of the surgery. But the downstream causal effects on hip rotation (body function) or overall gait pattern (GDI) involves possible patient compensations, rehabilitation details, and so on. Further supporting this conjecture is the fact that the largest observed MAE was for overall mobility as measured by the Functional Assessment Questionnaire Transform (FAQt). The FAQt measures activities, which are the most causally downstream outcome we predict.

**Table 2:**
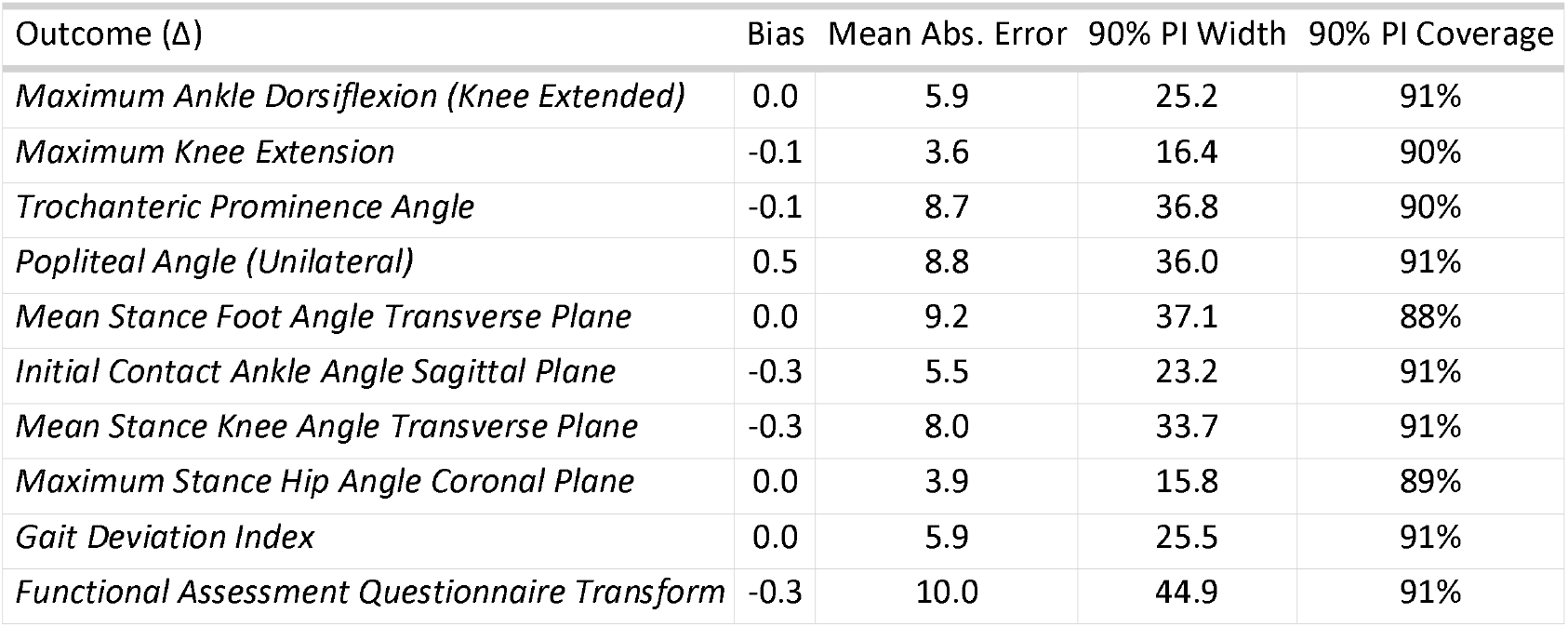
Outcome Model Performance.

#### Case Study Outcomes

We return to the case study introduced earlier. The patient underwent surgery at four levels during a single session, all on the right side. These were a femoral derotation osteotomy, hamstrings lengthening, calf muscle lengthening, and bony foot reconstruction. We note that the actual surgery was in concordance with the propensity models.

#### Single-Level Treatment

We can estimate the effect of specific treatments by setting those treatment variables to 1 while setting all other treatment variables to 0 in the outcome prediction models. We can also compute the predicted change in the outcome variable for an untreated limb by leaving all treatment variables at 0. The difference between these two predictions is the estimated treatment effect. We estimate the 50% and 90% prediction intervals for the treatment effect from the posterior distribution produced by the BART model.

In clinical practice, we generally report outcome predictions from each of 12 single-level surgeries. At the body structure level, most surgeries have isolated effects. For example, a femoral derotation osteotomy affects femoral torsion, but does not directly affect ankle dorsiflexion ROM. The reverse is true for a calf muscle lengthening. At the kinematic and activities levels, several surgeries can affect a single outcome. For example, both femoral and tibial derotation osteotomies may affect foot progression during gait. In situations like this, we generally recommend examining the predictions from each single-level surgery and summing the effects as an approximation. This approach makes the reasonable assumption that interactions are small.

#### Body Structure Level

For the femoral derotation osteotomy we estimated the change in femoral torsion. For the hamstrings lengthening we estimated the change in popliteal angle. For the calf muscle lengthening we estimated the change in passive ankle dorsiflexion ROM. For the bony foot surgery we estimate the change in overall weightbearing foot deformity severity [Figure 8]. We observe that the predicted outcomes are extremely accurate for change in ankle dorsiflexion ROM and anteversion, well within the 50% PI for change in weightbearing foot deformity, and at the edge of the 50% PI for change in popliteal angle. Note that by comparing the treated prediction (top, green range) to the untreated control prediction (bottom, purple range) we can estimate the *treatment effect* (middle, grey range). In this case, each surgery was predicted to have a relatively high probability of producing positive treatment effect at the body structures level. Keep in mind that treatment effect is an unobservable quantity, computed with the aid of an estimated counterfactual.

**Figure 8.**
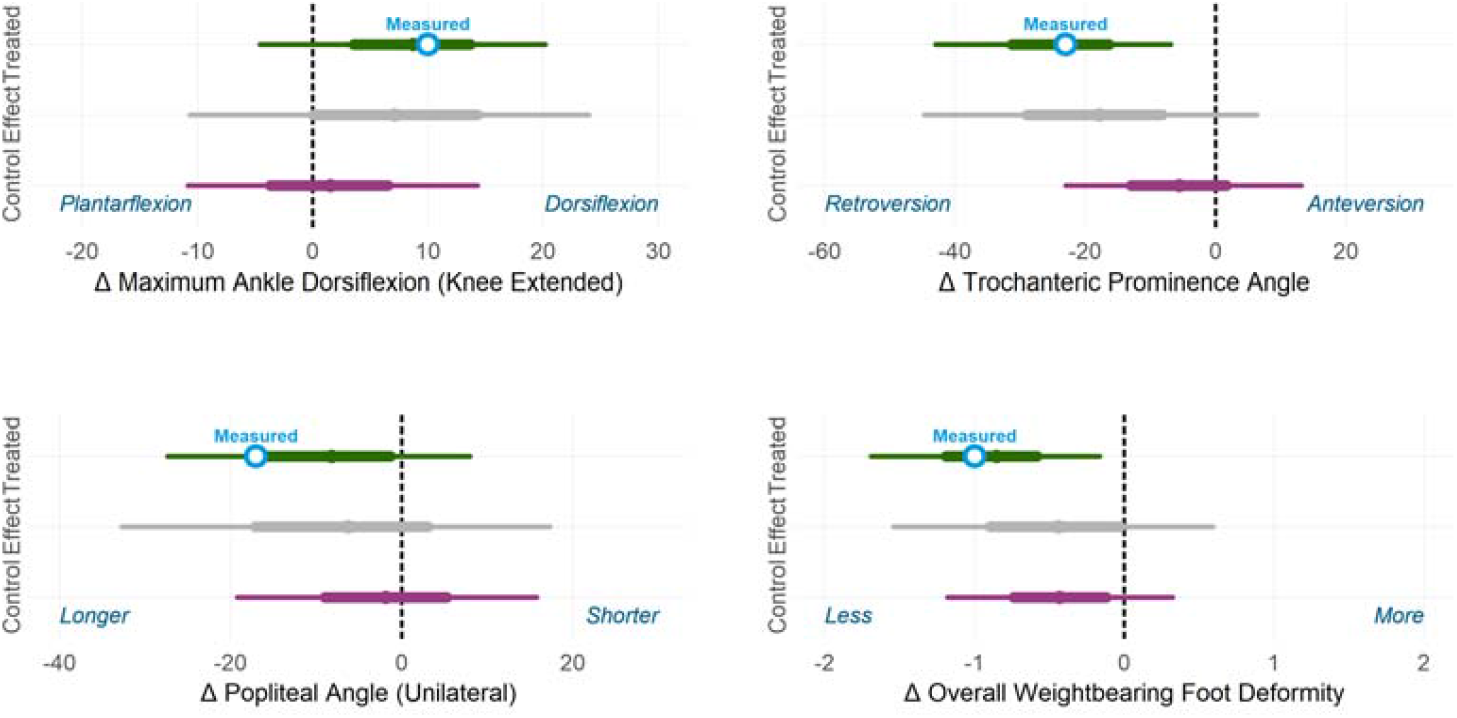
Body structure outcome. Measured outcomes are white circles. Prediction intervals are thick (50%) and thin (90%) lines, and a barely-visible solid point indicates the mean prediction.

#### Body Function Level (Kinematics)

We also estimated kinematic outcomes, which are at the level of body functions. For the femoral derotation osteotomy we estimated the change in hip rotation. For the hamstrings lengthening we estimated the change in initial contact knee flexion. For the calf muscle lengthening we estimated the change in initial contact ankle dorsiflexion. For the bony foot surgery we estimated the change in mean stance foot progression [Figure 9]. The measured changes in ankle dorsiflexion at initial contact, hip rotation during stance, and foot progression during stance were all within the 50% prediction interval, while the measured change in knee flexion at initial contact was larger than predicted, falling outside the 50% prediction interval but well within the 90% prediction interval. Note that this is sensible, given the slightly-larger-than-expected improvement in popliteal angle [Figure 8].

**Figure 9.**
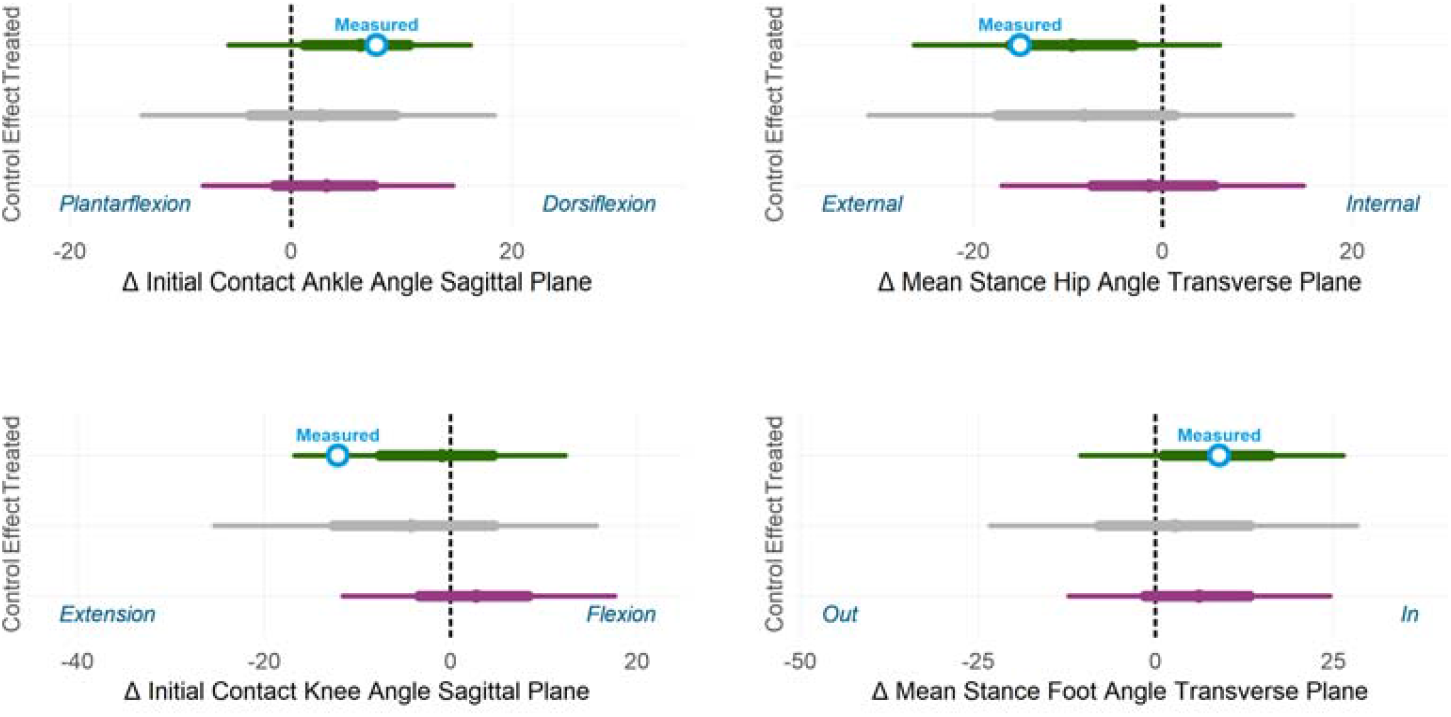
Body function (gait kinematics) outcome. Measured is white circle. Prediction intervals are thick (50%) and thin (90%) lines, and a small solid point indicates the mean prediction.

#### Multi-Level Treatment

In addition to modeling the outcomes of single-level treatment, the outcome prediction models can be used to estimate the effects of multi-level surgery. For the treated right limb of the case study, we estimated the structural and kinematic outcomes of interest for the multi-level surgery administered (femoral derotation osteotomy, hamstrings lengthening, calf muscle lengthening, and bony foot and ankle surgery). Estimated changes were all within the 50% prediction intervals [Figure 10]. The outcome estimates from multi-level surgery were similar to those obtained from single-level surgery, demonstrating — for this case — the claim that individual surgeries often affect specific body structures and functions with limited interactions.

**Figure 10.**
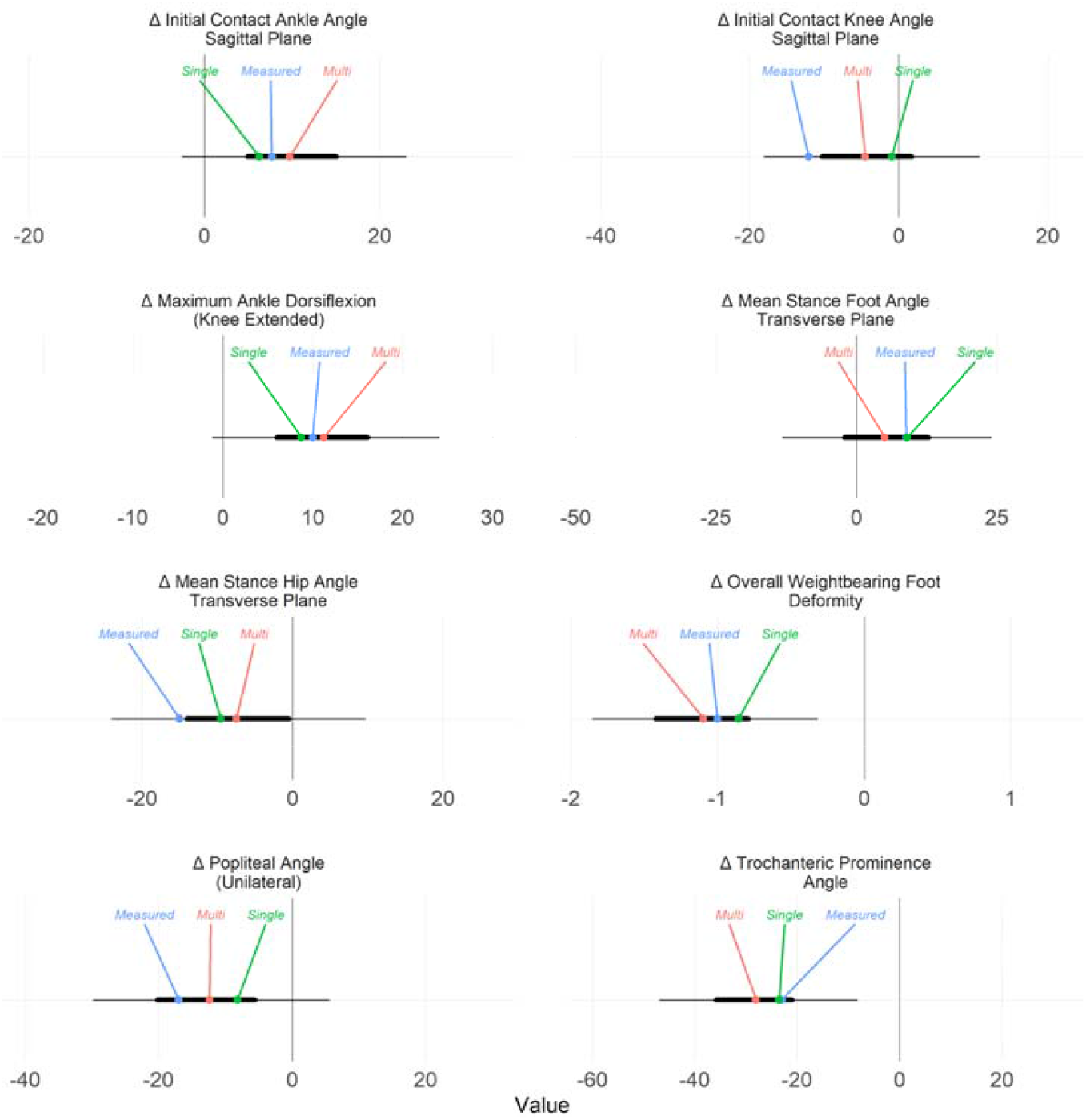
Multi-level (Multi) and single-level (Single) surgery predictions compared to measured outcomes for the treated side of the case-study individual. The multi- and single-level predictions are similar for all chosen outcomes.

### 2.4 Estimated Benefit of EB-GAIT

Using treatment recommendation and outcome prediction models represents a significant departure from the conventional approach to interpreting clinical gait data. At the outset of this manuscript, we stated that the motivation for a change was to improve the modest, stagnant, and unpredictable outcomes observed over the last several decades [Figure 1]. Therefore, it is important to estimate the potential benefit of adopting the EB-GAIT approach.

We can estimate the potential impact by simulating different ways the technique could be implemented: (1) *Continue current practice* - ignore the propensity and outcome models, and continue to make treatment decisions the way we have for over 30 years, (2) *Follow the propensity models only* - surgeries are administered if they have a probability greater than 50%, (3) *Follow the outcome models only* - surgeries are administered if their mean predicted outcome exceeds a threshold chosen by the panel of clinical experts, or (4) *Follow both the propensity and outcome models* - surgeries are administered if they have a probability greater than 50% and their mean predicted outcome exceeds a threshold chosen by the panel of clinical experts.

In practice, we do not recommend that propensity or outcome models should be blindly followed 100% of the time. There are countless factors affecting treatment decisions that are not captured by the models. The models are intended to provide supporting information to the treating clinician, not to replace their clinical judgment.

We examine the effects of these four implementations on overall kinematic outcomes (GDI) of limbs undergoing femoral derotation osteotomy [Figure 11]. A predicted GDI change of at least +7.5 was set as the threshold for a good outcome. The results indicate that relying solely on the propensity models has no significant impact on outcomes and leads to an increase in the number of limbs treated. Using only the outcome prediction models improves outcomes for treated limbs but slightly worsens outcomes for untreated limbs, while also resulting in a small increase in the number of limbs treated. In contrast, combining both the propensity and outcome models is estimated to enhance outcomes for treated limbs, maintain outcomes for untreated limbs, and substantially reduce the number of limbs treated. These findings suggest that adopting the EB-GAIT approach in the future could provide meaningful benefits.

**Figure 11.**
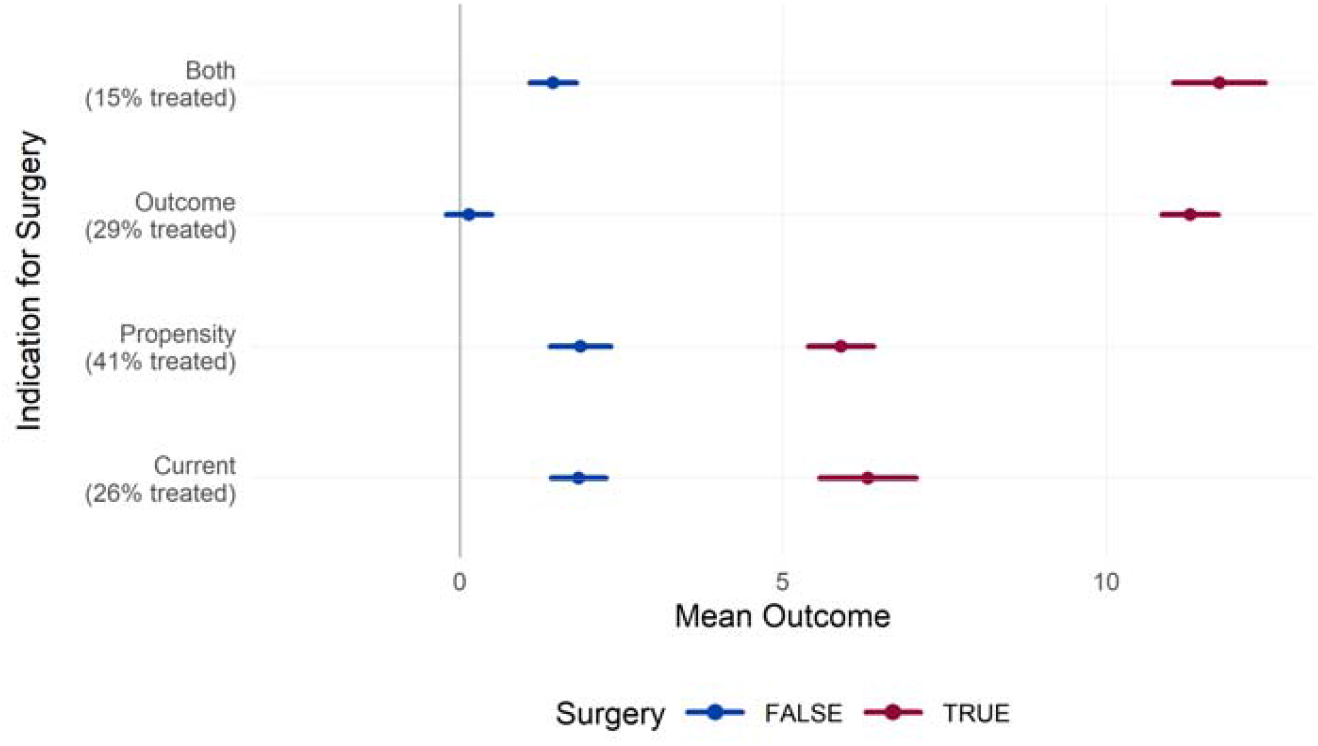
Change in GDI with- or without femoral derotation osteotomy. Following treatment recommendation and outcome prediction models leads to improved GDI on treated limbs, unchanged outcomes on untreated limbs and fewer limbs undergoing surgery. This suggests there can be a meaningful benefit to adopting the EB-GAIT approach in the future.

## 3. Discussion

We have introduced a novel approach for interpreting clinical gait data, termed the *Evidence-Based Gait Analysis Interpretation Technique* (EB-GAIT). At its core, this method leverages treatment recommendation and outcome prediction models to inform individualized treatment decisions. The treatment recommendation (propensity) models demonstrated high levels of balanced accuracy, sensitivity, specificity, and discriminatory power, while the outcome models were unbiased, provided reliable prediction intervals, and exhibited reasonable accuracy. The role of EB-GAIT is to estimate the likelihood of various treatments and their expected outcomes, while the clinician’s role is to thoughtfully integrate this information—along with their comprehensive understanding of the patient—into their decision-making process.

Our simulation of EB-GAIT in practice suggests that it has the potential to significantly improve upon the modest, stagnant, and unpredictable outcomes historically associated with conventional CGA interpretation methods. However, these findings are based on data from a single center, and results may vary at other institutions. We believe that pooling data from multiple centers with diverse treatment philosophies represents the most promising direction for future research. Notably, there is already at least one major project underway aimed at achieving this goal [25].

We also illustrated how these models can be applied in practice by demonstrating their use in a representative case study. This demonstration incorporated many of the graphical and textual report we currently employ to communicate EB-GAIT results to treating clinicians. The report is designed to deliver a clear, concise, and evidence-based summary of the available treatment options and their expected outcomes for a patient. A de-identified version of the EB Report for the case study is included as a supplement to this manuscript.

### 3.1 Treatment Recommendation Models

The propensity models demonstrated strong performance, with balanced accuracy, specificity, and sensitivity comparable to those of commonly used diagnostic tools for critical medical decisions. However, the effectiveness of these models is inherently constrained by the level of historical consistency among clinicians. For instance, at our center, the decision to perform psoas lengthening surgery varies significantly depending on the clinician, which limits the performance of a propensity model. Additionally, treatment philosophies differ not only within institutions but also between them [26]. As a result, models that work well for one center may not be suitable for another. In essence, the current propensity models serve as a second opinion, answering the question, “*What would Gillette do?*” for a specific patient. These models can also be valuable for trainees or clinicians new to a center who may not yet be familiar with the local treatment philosophy and standard of practice (SOP).

While using propensity models to identify a historical SOP is both sensible and powerful, it comes with significant responsibility. The modest, stagnant, and unpredictable historical outcomes suggest that the historical SOP may be suboptimal — potentially significantly so for certain treatments. Consequently, adhering strictly to a propensity model — even a perfect one — would be illogical. We encapsulate this idea with the mantra, “*Propensity is not Fate*.” The models presented here are designed to complement and enhance clinical expertise, not to replace it.

One safeguard against blindly following propensity models is the inclusion of model explanations based on Shapley values. These explanations provide clear insights into the factors driving the decision to recommend a specific treatment. If the clinician reviewing the EB-GAIT report disagrees with the reasoning behind the model’s prediction, they can choose to disregard or mentally adjust the recommendation. Additionally, the models provide a probability range, which helps quantify the uncertainty in the prediction. This range serves as a reminder that treatment decisions can vary due to differences in clinician judgment, as well as uncertainties inherent in the data and modeling process. By highlighting both the reasoning and the uncertainty, the EB-GAIT report encourages thoughtful, informed decision — making rather than uncritical adherence to model outputs.

### 3.2 Outcome Prediction Models

The outcome prediction models performed well, demonstrating unbiased results, reliable prediction intervals, and precision on par with experimental measurement error. These characteristics are encouraging. However, the prediction intervals were often wide — sometimes even comically so. While the models provide reasonable guidance on the general direction and magnitude of expected changes, reducing this uncertainty remains a key challenge. Identifying the sources of outcome variation among patients is both critical and daunting. “*Precision medicine*” is an aspirational goal rather than a current reality. In our deterministic universe, every observed outcome has underlying causes that, in theory, could be modeled. However, pinpointing those causes remains elusive.

Several strategies could help improve the outcome prediction models. These include enhancing the accuracy of input data, incorporating surgical details, adding additional model features, or exploring alternative modeling frameworks beyond BART. Addressing these areas could help narrow prediction intervals and improve the utility of the models in guiding clinical decision-making.

### 3.3 Implementation

An important consideration when using the EB-GAIT approach is the context in which it is applied. Currently, many clinical gait analysis practices involve treatment recommendations made by a physician who has neither examined nor met the patient in question. While we have demonstrated that, on a group level, EB-GAIT’s treatment recommendation and outcome prediction models can lead to better outcomes compared to traditional methods, the ultimate decision to pursue invasive treatments lies entirely with the patient and their medical care team. Therefore, we believe that EB-GAIT reports are most effectively utilized by the clinician who is considering offering the treatment, as they are intimately familiar with aspects of the patient that may not be captured in the evidence-based models.

The EB-GAIT report can be likened to an expert opinion provided to a treating clinician. It serves as a valuable piece of the patient care puzzle, offering evidence-based insights and recommendations. However, it is not absolute or infallible. The clinician’s expertise, combined with their knowledge of the patient’s unique circumstances, remains essential in making informed, personalized treatment decisions. The EB-GAIT report is designed to support, not replace, this critical clinical judgment.

The EB Report for the case discussed in this manuscript is included as a supplement. The report has been fully de-identified, including a synthetic name and record number, rounded age, truncated treatment event dates, and obscured identities in videos. It is a self-contained HTML file generated using the `Quarto` package from Posit [27]. In addition to the elements we have detailed extensively in this manuscript, the report includes background information, patient goals, observational videos, the complete physical examination, color coded to provide “*severity-at-a-glance*”, kinematic data, and graphs illustrating the progression of key measures over time.

The format of the EB Report is still evolving. The version provided here is current as of publication and represents a relatively mature and stable tool that we have been testing alongside our conventional methods for approximately one year. During this testing period, the report has been used as a supplement to — not a replacement for — the established conventional gait analysis interpretation method. We expect ongoing refinements to enhance the utility of these reports, driven by user feedback and further advancements in the field.

### 3.4 Conclusion

What we have described above represents a transformative approach to CGA, leveraging machine learning to provide data-driven treatment recommendations and outcome predictions. These tools are not intended to replace clinical judgment but to augment it, providing clinicians with guidance grounded in historical data and predictive analytics. Simulations suggest that adopting EB-GAIT could lead to improved outcomes, fewer unnecessary surgeries, and more consistent care. However, the utility of these models is inherently tied to the quality and consistency of historical data, and future efforts should focus on refining model inputs and pooling data from multiple centers to improve generalizability. The development of EB-GAIT marks a significant step toward precision medicine in gait analysis, offering a promising tool to enhance treatment outcomes and patient care.

## Data Availability

Data produced in the present study are available upon reasonable request to the authors

## Appendix 1 Treatment Recommendation Model Features

### Neural Rhizotomy

Era, Age, Diagnosis, Diagnosis Modifier, Adductor Spasticity, HamstringSpasticity, Plantarflexor Spasticity, Rectus Femoris Spasticity, PriorNeural Rhizotomy, Initial Contact Knee Angle Sagittal Plane, InitialContact Ankle Angle Sagittal Plane, Maximum Stance Ankle Angle SagittalPlane, Mean Swing Ankle Angle Sagittal Plane, GMFCS

### Rectus Transfer

Era, Age, Diagnosis, Diagnosis Modifier, Rectus Femoris Spasticity, PriorRectus Femoris Transfer, Prior Neural Rhizotomy, Initial Contact Knee AngleSagittal Plane, Maximum Swing Knee Angle Sagittal Plane

### Psoas Lengthening

Era, Age, Diagnosis, Diagnosis Modifier, Hip Flexor Strength, PoplitealAngle (Unilateral), Maximum Hip Extension, Prior Hams Lengthening, PriorNeural Rhizotomy, Mean Stance Pelvis Angle Sagittal Plane, Minimum StancePelvis Angle Sagittal Plane, Minimum Stance Hip Angle Sagittal Plane,Maximum Stance Pelvis Angle Sagittal Plane

### Hamstrings Lengthening

Era, Age, Diagnosis, Diagnosis Modifier, Maximum Knee Extension, PoplitealAngle (Unilateral), Prior Hams Lengthening, Prior Neural Rhizotomy, InitialContact Pelvis Angle Sagittal Plane, Initial Contact Knee Angle SagittalPlane, Mean Stance Knee Angle Sagittal Plane, Minimum Stance Knee AngleSagittal Plane, Mean Swing Pelvis Angle Sagittal Plane, Minimum SwingPelvis Angle Sagittal Plane, Maximum Swing Pelvis Angle Sagittal Plane

### Adductor Lengthening

Era, Age, Diagnosis, Diagnosis Modifier, Maximum Hip Abduction (KneeExtended), Maximum Hip Abduction (Knee Flexed), Hip Abductor Strength,Prior Adductor Release, Prior Neural Rhizotomy, Initial Contact Hip AngleCoronal Plane, Maximum Stance Hip Angle Coronal Plane, Minimum Swing HipAngle Coronal Plane

### Calf Muscle Lengthening

Era, Age, Diagnosis, Diagnosis Modifier, Maximum Ankle Dorsiflexion(Knee Extended), Maximum Ankle Dorsiflexion (Knee Flexed), MaximumKnee Extension, Plantarflexor Spasticity, Plantarflexor Strength,Non-Weightbearing Hindfoot Varus/Valgus, Non-Weightbearing HindfootVarus/Valgus Severity, Weightbearing Foot Position, WeightbearingFoot Position Severity, Prior Gastroc Soleus Lengthening, Prior NeuralRhizotomy, Initial Contact Knee Angle Sagittal Plane, Initial Contact AnkleAngle Sagittal Plane, Minimum Stance Ankle Angle Sagittal Plane, MaximumStance Ankle Angle Sagittal Plane, Minimum Swing Ankle Angle Sagittal Plane

### Femoral Derotation Osteotomy

Era, Age, Diagnosis, Diagnosis Modifier, EOS Femoral Anteversion, EOSBimalleolar Axis Angle, Trochanteric Prominence Angle, Bimalleolar AxisAngle, Maximum External Hip Rotation, Maximum Internal Hip Rotation,Weightbearing Forefoot Varus/Valgus, Weightbearing Forefoot Varus/ValgusSeverity, Weightbearing Forefoot Ab/Adduction, Weightbearing ForefootAb/Adduction Severity, Prior Tibial Derotation Osteotomy, Prior FemoralDerotation Osteotomy, Mean Stance Pelvis Angle Transverse Plane, MeanStance Hip Angle Transverse Plane, Mean Stance Knee Angle Transverse Plane,Mean Stance Foot Angle Transverse Plane

### Tibial Derotation Osteotomy

Era, Age, Diagnosis, Diagnosis Modifier, EOS Femoral Anteversion, EOSBimalleolar Axis Angle, Trochanteric Prominence Angle, Bimalleolar AxisAngle, Maximum External Hip Rotation, Maximum Internal Hip Rotation,Weightbearing Forefoot Varus/Valgus, Weightbearing Forefoot Varus/ValgusSeverity, Weightbearing Forefoot Ab/Adduction, Weightbearing ForefootAb/Adduction Severity, Prior Tibial Derotation Osteotomy, Prior FemoralDerotation Osteotomy, Mean Stance Pelvis Angle Transverse Plane, MeanStance Hip Angle Transverse Plane, Mean Stance Knee Angle Transverse Plane,Mean Stance Foot Angle Transverse Plane

### Distal Femoral Extension Osteotomy + Patellar Advancement

Era, Age, Diagnosis, Diagnosis Modifier, Extensor Lag, Maximum KneeExtension, Patella Alta, Prior Hams Lengthening, Prior DFEO, Prior PatellarAdvance, Mean Stance Knee Angle Sagittal Plane, Minimum Stance Knee AngleSagittal Plane

### Patellar Advancement

Era, Age, Diagnosis, Diagnosis Modifier, Extensor Lag, Maximum KneeExtension, Patella Alta, Prior Hams Lengthening, Prior DFEO, Prior PatellarAdvance, Mean Stance Knee Angle Sagittal Plane, Minimum Stance Knee AngleSagittal Plane

### Foot and Ankle Bony Reconstruction

Era, Age, Diagnosis, Diagnosis Modifier, Non-Weightbearing Arch,Non-Weightbearing Forefoot Varus/Valgus, Non-Weightbearing ForefootVarus/Valgus Severity, Non-Weightbearing Hindfoot Varus/Valgus,Non-Weightbearing Hindfoot Varus/Valgus Severity, Non-Weightbearing MidfootMotion, First Ray Plantarflexion, Weightbearing Forefoot Varus/Valgus,Weightbearing Forefoot Varus/Valgus Severity, Weightbearing ForefootAb/Adduction, Weightbearing Forefoot Ab/Adduction Severity, WeightbearingFoot Position, Weightbearing Foot Position Severity, Weightbearing MidfootPosition, Prior Foot and Ankle Bone, Prior Foot and Ankle Soft Tissue, MeanStance Foot Angle Transverse Plane

### Foot and Ankle Soft Tissue Reconstruction

Era, Age, Diagnosis, Diagnosis Modifier, Non-Weightbearing Arch,Non-Weightbearing Forefoot Varus/Valgus, Non-Weightbearing ForefootVarus/Valgus Severity, Non-Weightbearing Hindfoot Varus/Valgus,Non-Weightbearing Hindfoot Varus/Valgus Severity, Non-Weightbearing MidfootMotion, First Ray Plantarflexion, Weightbearing Forefoot Varus/Valgus,Weightbearing Forefoot Varus/Valgus Severity, Weightbearing ForefootAb/Adduction, Weightbearing Forefoot Ab/Adduction Severity, WeightbearingFoot Position, Weightbearing Foot Position Severity, Weightbearing MidfootPosition, Prior Foot and Ankle Bone, Prior Foot and Ankle Soft Tissue, MeanStance Foot Angle Transverse Plane, Mean Swing Foot Angle Transverse Plane

## Appendix 2 Outcome Prediction Model Features

### Maximum Ankle Dorsiflexion (Knee Extended)

Age, Maximum Ankle Dorsiflexion (Knee Extended), Maximum Ankle Dorsiflexion(Knee Flexed), Initial Contact Knee Angle Sagittal Plane, InitialContact Ankle Angle Sagittal Plane, Minimum Stance Knee Angle SagittalPlane, Maximum Stance Ankle Angle Sagittal Plane, Minimum Swing AnkleAngle Sagittal Plane, GMFCS, Interval Adductor Release, Interval DFEO +Patellar Advance, Interval Femoral Derotation Osteotomy, Interval Foot andAnkle Bone, Interval Foot and Ankle Soft Tissue, Interval Gastroc SoleusLengthening, Interval Hams Lengthening, Interval Neural Rhizotomy, IntervalPatellar Advance, Interval Psoas Release, Interval Rectus Femoris Transfer,Interval Tibial Derotation Osteotomy

### Maximum Knee Extension

Age, Extensor Lag, Maximum Knee Extension, Patella Alta, Mean Stance KneeAngle Sagittal Plane, Minimum Stance Knee Angle Sagittal Plane, GMFCS,Interval Adductor Release, Interval DFEO + Patellar Advance, IntervalFemoral Derotation Osteotomy, Interval Foot and Ankle Bone, Interval Footand Ankle Soft Tissue, Interval Gastroc Soleus Lengthening, Interval HamsLengthening, Interval Neural Rhizotomy, Interval Patellar Advance, IntervalPsoas Release, Interval Rectus Femoris Transfer, Interval Tibial DerotationOsteotomy

### Trochanteric Prominence Angle

Age, EOS Femoral Anteversion, EOS Bimalleolar Axis Angle, TrochantericProminence Angle, Bimalleolar Axis Angle, Maximum External Hip Rotation,Maximum Internal Hip Rotation, Mean Stance Pelvis Angle Transverse Plane,Mean Stance Hip Angle Transverse Plane, Mean Stance Knee Angle TransversePlane, Mean Stance Foot Angle Transverse Plane, GMFCS, Interval AdductorRelease, Interval DFEO + Patellar Advance, Interval Femoral DerotationOsteotomy, Interval Foot and Ankle Bone, Interval Foot and Ankle SoftTissue, Interval Gastroc Soleus Lengthening, Interval Hams Lengthening,Interval Neural Rhizotomy, Interval Patellar Advance, Interval PsoasRelease, Interval Rectus Femoris Transfer, Interval Tibial DerotationOsteotomy

### Popliteal Angle (Unilateral)

Age, Popliteal Angle (Unilateral), Initial Contact Pelvis Angle SagittalPlane, Initial Contact Knee Angle Sagittal Plane, Mean Stance Knee AngleSagittal Plane, Minimum Stance Knee Angle Sagittal Plane, GMFCS, IntervalAdductor Release, Interval DFEO + Patellar Advance, Interval FemoralDerotation Osteotomy, Interval Foot and Ankle Bone, Interval Foot andAnkle Soft Tissue, Interval Gastroc Soleus Lengthening, Interval HamsLengthening, Interval Neural Rhizotomy, Interval Patellar Advance, IntervalPsoas Release, Interval Rectus Femoris Transfer, Interval Tibial DerotationOsteotomy

### Mean Stance Foot Angle - Transverse Plane

Age, EOS Femoral Anteversion, EOS Bimalleolar Axis Angle, TrochantericProminence Angle, Bimalleolar Axis Angle, Maximum External Hip Rotation,Maximum Internal Hip Rotation, Mean Stance Pelvis Angle Transverse Plane,Mean Stance Hip Angle Transverse Plane, Mean Stance Knee Angle TransversePlane, Mean Stance Foot Angle Transverse Plane, GMFCS, Interval AdductorRelease, Interval DFEO + Patellar Advance, Interval Femoral DerotationOsteotomy, Interval Foot and Ankle Bone, Interval Foot and Ankle SoftTissue, Interval Gastroc Soleus Lengthening, Interval Hams Lengthening,Interval Neural Rhizotomy, Interval Patellar Advance, Interval PsoasRelease, Interval Rectus Femoris Transfer, Interval Tibial DerotationOsteotomy

### Initial Contact Ankle Angle - Sagittal Plane

Age, Maximum Ankle Dorsiflexion (Knee Extended), Maximum Ankle Dorsiflexion(Knee Flexed), Initial Contact Knee Angle Sagittal Plane, Initial ContactAnkle Angle Sagittal Plane, Mid-Stance Ankle Angle Sagittal Plane, MinimumStance Knee Angle Sagittal Plane, Minimum Swing Ankle Angle SagittalPlane, GMFCS, Interval Adductor Release, Interval DFEO + Patellar Advance,Interval Femoral Derotation Osteotomy, Interval Foot and Ankle Bone,Interval Foot and Ankle Soft Tissue, Interval Gastroc Soleus Lengthening,Interval Hams Lengthening, Interval Neural Rhizotomy, Interval PatellarAdvance, Interval Psoas Release, Interval Rectus Femoris Transfer, IntervalTibial Derotation Osteotomy

### Mean Stance Knee Angle - Transverse Plane

Age, EOS Femoral Anteversion, EOS Bimalleolar Axis Angle, TrochantericProminence Angle, Bimalleolar Axis Angle, Maximum External Hip Rotation,Maximum Internal Hip Rotation, Mean Stance Pelvis Angle Transverse Plane,Mean Stance Hip Angle Transverse Plane, Mean Stance Knee Angle TransversePlane, Mean Stance Foot Angle Transverse Plane, GMFCS, Interval AdductorRelease, Interval DFEO + Patellar Advance, Interval Femoral DerotationOsteotomy, Interval Foot and Ankle Bone, Interval Foot and Ankle SoftTissue, Interval Gastroc Soleus Lengthening, Interval Hams Lengthening,Interval Neural Rhizotomy, Interval Patellar Advance, Interval PsoasRelease, Interval Rectus Femoris Transfer, Interval Tibial DerotationOsteotomy

### Max. Stance Hip Angle - Coronal Plane

Age, Maximum Hip Abduction (Knee Extended), Initial Contact Hip AngleCoronal Plane, Maximum Stance Hip Angle Coronal Plane, Minimum Swing HipAngle Coronal Plane, GMFCS, Interval Adductor Release, Interval DFEO +Patellar Advance, Interval Femoral Derotation Osteotomy, Interval Foot andAnkle Bone, Interval Foot and Ankle Soft Tissue, Interval Gastroc SoleusLengthening, Interval Hams Lengthening, Interval Neural Rhizotomy, IntervalPatellar Advance, Interval Psoas Release, Interval Rectus Femoris Transfer,Interval Tibial Derotation Osteotomy

### Gait Deviation Index

Overall Spasticity, Overall Weightbearing Foot Deformity, OverallNon-Weightbearing Foot Deformity, Age, EOS Femoral Anteversion, EOSBimalleolar Axis Angle, Maximum Ankle Dorsiflexion (Knee Extended),Maximum Ankle Dorsiflexion (Knee Flexed), Trochanteric Prominence Angle,Bimalleolar Axis Angle, Extensor Lag, Maximum External Hip Rotation,Maximum Internal Hip Rotation, Maximum Knee Extension, Patella Alta,Popliteal Angle (Unilateral), Gait Deviation Index, Maximum Hip Extension,Initial Contact Pelvis Angle Coronal Plane, Initial Contact Pelvis AngleSagittal Plane, Initial Contact Pelvis Angle Transverse Plane, InitialContact Hip Angle Coronal Plane, Initial Contact Hip Angle Sagittal Plane,Initial Contact Hip Angle Transverse Plane, Initial Contact Knee AngleSagittal Plane, Initial Contact Ankle Angle Sagittal Plane, Initial ContactFoot Angle Transverse Plane, Opposite Foot Off Pelvis Angle Coronal Plane,Opposite Foot Off Pelvis Angle Sagittal Plane, Opposite Foot Off PelvisAngle Transverse Plane, Opposite Foot Off Hip Angle Coronal Plane, OppositeFoot Off Hip Angle Sagittal Plane, Opposite Foot Off Hip Angle TransversePlane, Opposite Foot Off Knee Angle Sagittal Plane, Opposite Foot OffAnkle Angle Sagittal Plane, Opposite Foot Off Foot Angle Transverse Plane,Opposite Foot Contact Pelvis Angle Coronal Plane, Opposite Foot ContactPelvis Angle Sagittal Plane, Opposite Foot Contact Pelvis Angle TransversePlane, Opposite Foot Contact Hip Angle Coronal Plane, Opposite Foot ContactHip Angle Sagittal Plane, Opposite Foot Contact Hip Angle Transverse Plane,Opposite Foot Contact Knee Angle Sagittal Plane, Opposite Foot ContactAnkle Angle Sagittal Plane, Opposite Foot Contact Foot Angle TransversePlane, Foot Off Pelvis Angle Coronal Plane, Foot Off Pelvis Angle SagittalPlane, Foot Off Pelvis Angle Transverse Plane, Foot Off Hip Angle CoronalPlane, Foot Off Hip Angle Sagittal Plane, Foot Off Hip Angle TransversePlane, Foot Off Knee Angle Sagittal Plane, Foot Off Ankle Angle SagittalPlane, Foot Off Foot Angle Transverse Plane, Mid-Swing Pelvis AngleCoronal Plane, Mid-Swing Pelvis Angle Sagittal Plane, Mid-Swing PelvisAngle Transverse Plane, Mid-Swing Hip Angle Coronal Plane, Mid-Swing HipAngle Sagittal Plane, Mid-Swing Hip Angle Transverse Plane, Mid-Swing KneeAngle Sagittal Plane, Mid-Swing Ankle Angle Sagittal Plane, Mid-Swing FootAngle Transverse Plane, GMFCS, Interval Adductor Release, Interval DFEO +Patellar Advance, Interval Femoral Derotation Osteotomy, Interval Foot andAnkle Bone, Interval Foot and Ankle Soft Tissue, Interval Gastroc SoleusLengthening, Interval Hams Lengthening, Interval Neural Rhizotomy, IntervalPatellar Advance, Interval Psoas Release, Interval Rectus Femoris Transfer,Interval Tibial Derotation Osteotomy

### Functional Assessment Questionnaire Transform

Overall Spasticity, Overall Weightbearing Foot Deformity, OverallNon-Weightbearing Foot Deformity, Age, Functional Assessment QuestionnaireTransform, GMFCS, Interval Adductor Release, Interval DFEO + PatellarAdvance, Interval Femoral Derotation Osteotomy, Interval Foot and AnkleBone, Interval Foot and Ankle Soft Tissue, Interval Gastroc SoleusLengthening, Interval Hams Lengthening, Interval Neural Rhizotomy, IntervalPatellar Advance, Interval Psoas Release, Interval Rectus Femoris Transfer,Interval Tibial Derotation Osteotomy

## Notes

Treatment Recommendation and Outcome Prediction Models to Support Decision-Making Based on Clinical Gait Analysis Data

### Competing Interest Statement

The authors have declared no competing interest.

### Funding Statement

This study did not receive any funding

### Author Declarations

This study was reviewed and authorized by the University of Minnesota institutional review board review (STUDY00012420). All experiments were performed in accordance with relevant guidelines and regulations. Informed consent for use of medical records was obtained at the time of service from all participants or their legal guardian. An option to rescind this permission is offered to patients at every visit to our center.

